# Fixed-Dose Combination Use and Drug Adherence, Healthcare Utilization, and Costs

**DOI:** 10.1101/2024.11.27.24318118

**Authors:** Donglan Zhang, Jun Soo Lee, Nicole L. Therrien, Lisa M. Pollack, Sandra L. Jackson, Xiaobei Dong, Anand Rajan, Kai Hong, Andrew E. Moran, Feijun Luo

## Abstract

**BACKGROUND:** Combination-pill therapy using fixed-dose combination (FDC) antihypertensives has the potential to improve hypertension control and management. However, combination-pill therapy remains low. This study aims to assess 1) the association between combination-pill therapy and subsequent medication adherence, healthcare utilization, and costs and 2) the potential to mitigate racial and ethnic disparities in medication adherence, among Medicaid beneficiaries.

**METHODS:** A retrospective cohort analysis was conducted using the 2017-2021 Merative^TM^ MarketScan® Medicaid claims database. The study sample included adults aged 18-64 years with hypertension, continuously enrolled one year before and after a random index date of prescribed antihypertensives. Combination-pill therapy was compared with multi-pill therapy. The propensity-score overlap weighting method was used to balance characteristics between individuals using combination- and multi-pill therapy. Logistic models were used for medication adherence, linear models for medication possession ratios (MPRs), negative binomial models for healthcare utilization, and generalized linear models for healthcare costs.

**RESULTS:** Compared with multi-pill therapy, combination-pill therapy was associated with improved medication adherence (3.17 in MPR, 95% CI: 2.79 - 3.55), fewer hypertension-related emergency department visits (220 per 1,000 individuals, 95% CI: -235 – -204), fewer hospitalizations (153 per 1,000 individuals, 95% CI: -160 – -146), and lower medical costs ($2,862 per person, 95% CI: -$3,035 – -$2,689). However, disparities in medication adherence between non-Hispanic White and non-Hispanic Black adults existed for both FDC and multi-pill therapy.

**CONCLUSIONS:** Combination-pill therapy could improve hypertension management and save costs for the Medicaid program and beneficiaries. These findings may inform policies on FDC antihypertensive coverage and programs.

Disclaimer: The findings and conclusions in this report are those of the authors and do not necessarily represent the official position of the Centers for Disease Control and Prevention. Use of trade names and commercial sources is for identification only and does not imply endorsement by the U.S. Department of Health and Human Services.

## Introduction

Hypertension is a major risk factor for cardiovascular disease, stroke, kidney disease, and dementia,^1–3^ affecting about half of Americans based on the 2017 American College of Cardiology and American Heart Association (ACC/AHA) definition for hypertension (threshold: 130/80 mmHg).^4^ Non-Hispanic Black (NHB) adults have substantially higher hypertension prevalence but lower medication adherence and prevalence of blood pressure control than Non-Hispanic White (NHW) and Hispanic adults.^5,6^

Fixed-dose combination (FDC) therapy that combines ≥2 antihypertensive medications into a single pill (a treatment strategy referred to as combination-pill therapy in this study) simplifies medication regimens and has been proven safe and effective.^7^ Combination-pill therapy using FDC antihypertensives may improve medication adherence and hypertension control.^8^ The 2017 ACC/AHA guideline recommended lower blood pressure thresholds and using combination-pill antihypertensive therapy as an initial management strategy for some individuals to control hypertension more quickly and effectively.^9^ The 2018 European Society of Cardiology/European Society of Hypertension and the Japanese Society of Hypertension guidelines also highlighted the importance of FDC-antihypertensives for hypertension treatment and recommend initial combination-pill therapy in almost all patients.^10^

Despite the promise of combination-pill therapy, its utilization for hypertension treatment in the United States remains low.^11,12^ This could be due to several reasons. One is that certain FDC-antihypertensives, although approved, may not be included in health care systems’ treatment protocols and insurance coverage formularies.^13,14^ Therapeutic inertia may also play a role, and there are concerns among health care professionals regarding the safety and efficacy, availability of different combinations, insurance coverage, and access to detailed FDC-antihypertensive information.^15^ These barriers may particularly affect low-income and racial and ethnic minority populations, who may disproportionately face financial burden if insurance does not cover these drugs.^16,17^ Evidence is lacking regarding whether combination-pill therapy is effective in mitigating racial and ethnic disparities in hypertension management.

To address this evidence gap, using large-scale claims data, we conducted an analysis among Medicaid beneficiaries to investigate: 1) the association between combination-pill therapy and subsequent medication adherence, healthcare utilization and medical expenses over a one-year period, and 2) whether the associations between combination-pill therapy and hypertension-related outcomes vary across racial and ethnic groups.

## Methods

### Data Sources and Study Population

Data were obtained from the Merative^TM^ MarketScan^®^ multistate Medicaid claims database from 2016 to 2022. To build our study sample (**Appendix Figure 1**), we first identified Medicaid beneficiaries who had ≥1 hypertension diagnosis (ICD-10-CM codes I10-I15) and had ≥1 pharmacy claims for an antihypertensive medication between January 2017 and December 2021 (study period, **Appendix Table 1**). During this period, we selected a random date as the index date to define the time when individuals filled or refilled antihypertensive medications. The random selection of the index date is preferred over identifying an index date based on the order of a pharmacy claim, as the latter would oversample individuals from earlier years compared with later years.^18^ A random index date allows for a more even distribution of individuals who initiated medications at different times across the study period and has been widely used in claims data analysis.^18,19^ We then restricted the sample to those who (1) were aged 18-64 years at the index date, (2) were continuously enrolled in one Medicaid insurance plan for the entire 1-year follow-up period post-index date, (3) had one year of data for prescribed antihypertensives before and after the index date.

We excluded beneficiaries aged ≥65 years who may have been dually eligible for Medicare, as well as those who had a pregnancy diagnosis during the study period, as pregnancy may affect medication regimens.^20^ In addition, to streamline the comparison between those using combination-pill therapy and those using non-FDC-antihypertensives for two different medication classes (referred to as multi-pill therapy in this study), we excluded beneficiaries who switched between combination-pill therapy and multi-pill therapy. We also excluded those who used only one antihypertensive therapeutic class to create comparable groups for the two therapy types. Finally, when calculating the annual healthcare costs, we identified a subset of adults not covered by capitated health insurance plans because fixed annual payment model plans do not fully reflect healthcare costs incurred when receiving services.^21^

### Outcomes

The primary outcomes of the study included 1) antihypertensive medication adherence within one year post-index date, 2) emergency department (ED) visits and inpatient admissions associated with hypertension and cardiovascular disease (CVD) per 1,000 individuals within one-year post-index date, and 3) hypertension-related medical costs (overall, and separately for ED visits and inpatient admissions) within one year post-index date and 4) CVD-related medical costs (overall, and separately for ED visits and inpatient admissions) within one year post-index date. Sensitivity analyses examined all-cause costs. Hypertension-related and CVD-related healthcare utilization and total medical costs were defined based on the presence of the respective diagnosis codes (hypertension: ICD-10-CM codes I10-I15; CVD: ICD-10-CM codes I00-I78).^22^

Medication adherence was measured using the medication possession ratio (MPR), with MPR ≥80% indicating adequate adherence to antihypertensive medications.^23,24^ The average MPR was computed as the ratio of the total days of antihypertensive drug supply to the number of days in each year.^23^ For combination-pill therapy, we derived the average MPRs for all available FDC-antihypertensives in the database (**Appendix Table 2**) and for the seven antihypertensive therapeutic classes including Angiotensin-Converting Enzyme (ACE) inhibitors, angiotensin receptor blockers (ARB), beta-blockers, calcium channel blockers (CCB), diuretics, other antihypertensives, and renin-angiotensin system antagonists separately, along with the corresponding generic drug names.^24^ For multi-pill therapy, we derived the average MPRs for each antihypertensive medication.

For our secondary outcomes, we assessed 1) Medicaid insurance payments (including all adjusted payments for healthcare services from the Medicaid program), 2) patient out-of-pocket (OOP) costs (including copay, coinsurance and deductibles) and 3) total costs (the sum of the Medicaid insurance payments and patient OOP costs for combination-and multi-pill therapy per individual. All secondary outcomes reflect results within one-year post-index date.

All outcomes were stratified by race and ethnicity (NHW, NHB, and Hispanic populations).

We did not stratify by other racial groups due to their high heterogeneity.

### Exposure

Combination-pill therapy was defined as having a pharmacy claim for ≥1 FDC-antihypertensive medication and no other antihypertensive medications from the index date through the follow-up period. The comparison was multi-pill therapy, defined as having no claims for FDC-antihypertensives but having claims for ≥2 different antihypertensive medication classes from the index date through the follow-up period.

### Statistical Analysis

We used propensity score (PS) overlap weighting methods to balance characteristics between individuals using combination- and multi-pill therapy (**Appendix Figure 2**).^25,26^ In calculating the PS, we included age groups (18-34, 35-44, 45-54, and 55-64 years), gender, race and ethnicity (NHW, NHB, Hispanic, and other racial groups), dummy indicators for risk factors (alcohol use, tobacco use, obesity, and lipid disorders), and 12 comorbidities identified in the Charlson comorbidity index (CCI, excluding myocardial infarction, congestive heart failure, peripheral vascular disease, cerebrovascular disease, and renal disease from the original CCI because these comorbidities may be in the causal pathways between antihypertensive medication use and CVD outcomes).^27^. All sociodemographic and comorbidity variables were measured in the one-year period prior to the index date.

We employed logistic models for medication adherence and linear models for MPRs. We then employed negative binomial models to estimate all healthcare utilization outcomes. We utilized generalized linear models with gamma distribution and log link function to accommodate the skewed distribution of healthcare costs.^28^ In sensitivity analyses, we included those who used both combination-pill *and* multi-pill therapy in the same year (to create a more inclusive definition of the combination-pill therapy group, allowing for inclusion in the exposure group of patients who may have switched between regimens or who may have been taking an FDC-antihypertensive along with additional antihypertensive agents simultaneously) and tested the robustness of the results.

All models were additionally adjusted for fixed effects for the year and month of the index date to control for time-invariant fixed effects within a year or a month. In each model, we compared outcomes between combination-and multi-pill therapy groups within the entire population, as well as within each race and ethnicity. We tested whether outcomes between combination- and multi-pill therapy groups differed by race/ethnicity by examining the statistical significance of interaction terms between therapy type and race/ethnicity.

Average marginal effects and corresponding 95% confidence intervals (95% CI) were reported. Sensitivity analyses were conducted to assess the association between combination-pill therapy and all-cause healthcare utilization, as combination-pill therapy may have spillover effects on other disease outcomes and related healthcare utilization. All analyses were conducted using Stata SE 17.0 statistical software (StataCorp, College Station, TX) and SAS version 9.4 (SAS 9.4, Cary, NC. SAS Institute Inc.).

## Results

After sample selection, a total of 325,600 adults with Medicaid coverage were included in the analysis of healthcare utilization outcomes. In the subset analysis among those not covered by capitated insurance, 128,528 adults were included in the assessment of cost outcomes (**Appendix Figure 1**).

### Sample Characteristics

Among 325,600 adults, 29,328 (9%) used combination-pill therapy to manage their hypertension, while 296,272 (91%) used multi-pill therapy (**Table 1**). Compared with the multi-pill therapy group, the combination-pill therapy group was significantly younger (48.2 vs. 49.4, p<0.001) and had a higher proportion of females (64.11% vs. 55.01%, p<0.001) and NHB adults (33.12% vs. 28.15%, p<0.001). The combination-pill therapy group also had a lower proportion of alcohol and tobacco use (both p<0.001) and a lower proportion of some comorbidities (dementia, chronic pulmonary disease, rheumatic disease and diabetes), but higher proportion of others (obesity, diabetes, and lipid disorders) (all p<0.001). After weighting the sample using the PS overlap approach, the characteristics between individuals using combination-pill and multi-pill therapy were not statistically significant (**Appendix Figure 2**).

**Table 1.**
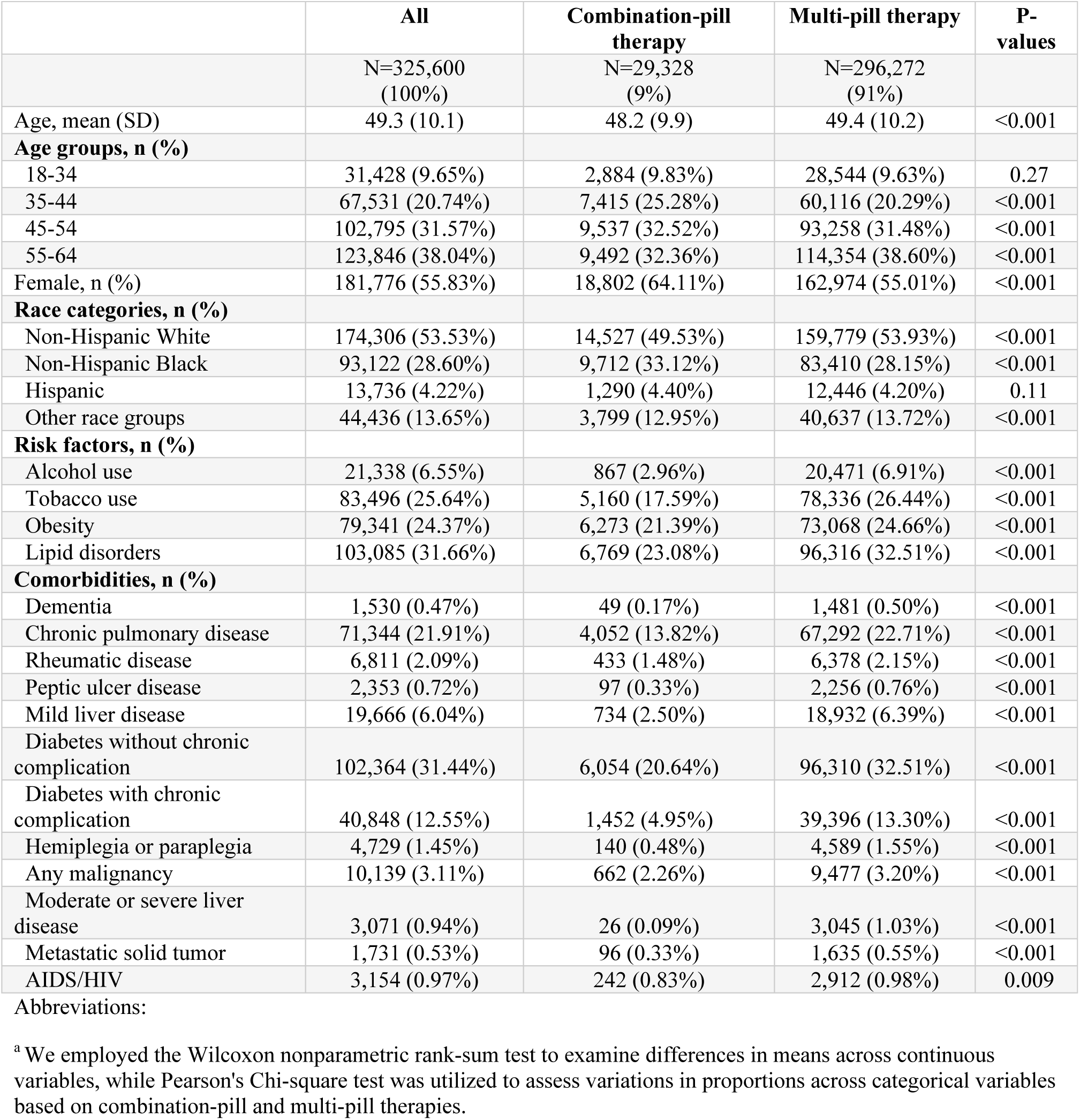
Summary Statistics of Sample Characteristics

|  | All | Combination-pill therapy | Multi-pill therapy | P-values |
| --- | --- | --- | --- | --- |
|  | N=325,600 (100%) | N=29,328 (9%) | N=296,272 (91%) |  |
| Age, mean (SD) | 49.3 (10.1) | 48.2 (9.9) | 49.4 (10.2) | <0.001 |
| **Age groups, n (%)** |  |  |  |  |
| 18-34 | 31,428 (9.65%) | 2,884 (9.83%) | 28,544 (9.63%) | 0.27 |
| 35-44 | 67,531 (20.74%) | 7,415 (25.28%) | 60,116 (20.29%) | <0.001 |
| 45-54 | 102,795 (31.57%) | 9,537 (32.52%) | 93,258 (31.48%) | <0.001 |
| 55-64 | 123,846 (38.04%) | 9,492 (32.36%) | 114,354 (38.60%) | <0.001 |
| Female, n (%) | 181,776 (55.83%) | 18,802 (64.11%) | 162,974 (55.01%) | <0.001 |
| **Race categories, n (%)** |  |  |  |  |
| Non-Hispanic White | 174,306 (53.53%) | 14,527 (49.53%) | 159,779 (53.93%) | <0.001 |
| Non-Hispanic Black | 93,122 (28.60%) | 9,712 (33.12%) | 83,410 (28.15%) | <0.001 |
| Hispanic | 13,736 (4.22%) | 1,290 (4.40%) | 12,446 (4.20%) | 0.11 |
| Other race groups | 44,436 (13.65%) | 3,799 (12.95%) | 40,637 (13.72%) | <0.001 |
| **Risk factors, n (%)** |  |  |  |  |
| Alcohol use | 21,338 (6.55%) | 867 (2.96%) | 20,471 (6.91%) | <0.001 |
| Tobacco use | 83,496 (25.64%) | 5,160 (17.59%) | 78,336 (26.44%) | <0.001 |
| Obesity | 79,341 (24.37%) | 6,273 (21.39%) | 73,068 (24.66%) | <0.001 |
| Lipid disorders | 103,085 (31.66%) | 6,769 (23.08%) | 96,316 (32.51%) | <0.001 |
| **Comorbidities, n (%)** |  |  |  |  |
| Dementia | 1,530 (0.47%) | 49 (0.17%) | 1,481 (0.50%) | <0.001 |
| Chronic pulmonary disease | 71,344 (21.91%) | 4,052 (13.82%) | 67,292 (22.71%) | <0.001 |
| Rheumatic disease | 6,811 (2.09%) | 433 (1.48%) | 6,378 (2.15%) | <0.001 |
| Peptic ulcer disease | 2,353 (0.72%) | 97 (0.33%) | 2,256 (0.76%) | <0.001 |
| Mild liver disease | 19,666 (6.04%) | 734 (2.50%) | 18,932 (6.39%) | <0.001 |
| Diabetes without chronic complication | 102,364 (31.44%) | 6,054 (20.64%) | 96,310 (32.51%) | <0.001 |
| Diabetes with chronic complication | 40,848 (12.55%) | 1,452 (4.95%) | 39,396 (13.30%) | <0.001 |
| Hemiplegia or paraplegia | 4,729 (1.45%) | 140 (0.48%) | 4,589 (1.55%) | <0.001 |
| Any malignancy | 10,139 (3.11%) | 662 (2.26%) | 9,477 (3.20%) | <0.001 |
| Moderate or severe liver disease | 3,071 (0.94%) | 26 (0.09%) | 3,045 (1.03%) | <0.001 |
| Metastatic solid tumor | 1,731 (0.53%) | 96 (0.33%) | 1,635 (0.55%) | <0.001 |
| AIDS/HIV | 3,154 (0.97%) | 242 (0.83%) | 2,912 (0.98%) | 0.009 |

Abbreviations:
<sup>a</sup> We employed the Wilcoxon nonparametric rank-sum test to examine differences in means across continuous variables, while Pearson's Chi-square test was utilized to assess variations in proportions across categorical variables based on combination-pill and multi-pill therapies.

### Association between Combination-Pill Therapy and Medication Adherence

After PS overlap weighting, compared with multi-pill therapy, combination-pill therapy was associated with, on average, an 8.5 percentage point (95% CI: 7.9 – 9.1) higher likelihood of medication adherence (MPR ≥ 80%) or 3.17 (95% CI: 2.79 – 3.55) higher average MPR (**Table 2**). The associations were strongest among NHW adults, who had a 9.7 percentage point (95% CI: 8.9 – 10.5) higher likelihood of medication adherence when using combination-pill vs multi-pill therapy, compared with the 6.9 percentage point (95% CI: 5.9 – 7.9) higher likelihood among NHB and 5.7 percentage point (95% CI: 2.9 – 8.5) among Hispanic adults (p<0.001).

**Table 2.** The Association of Combination-Pill Therapy Use with Medication Adherence and Medication Possession Ratios (per individual)^a^

|  | All | NH White | NH Black | Hispanic |
| --- | --- | --- | --- | --- |
| **Medication Adherence (MPR ≥ 80%) to Antihypertensives** |  |  |  |  |
| Multi-pill therapy<sup>b</sup> | 0.395<br>(0.393 - 0.397) | 0.444<br>(0.442 - 0.447) | 0.309<br>(0.305 - 0.312) | 0.386<br>(0.376 - 0.395) |
| Combination-pill therapy only<sup>b</sup> | 0.480<br>(0.474 - 0.485) | 0.541<br>(0.534 - 0.549) | 0.378<br>(0.368 - 0.387) | 0.442<br>(0.416 - 0.469) |
| Difference<sup>c</sup> | **0.085\*\*\***<br>**(0.079 - 0.091)** | **0.097\*\*\***<br>**(0.089 - 0.105)** | **0.069\*\*\***<br>**(0.059 - 0.079)** | **0.057\*\*\***<br>**(0.029 - 0.085)** |
| Observations | 325,600 | 174,306 | 93,122 | 13,736 |
| **Medication Possession Ratios** |  |  |  |  |
| Multi-pill therapy | 65.66<br>(65.53 - 65.78) | 69.23<br>(69.06 - 69.40) | 59.27<br>(59.04 - 59.50) | 66.33<br>(65.72 - 66.93) |
| Combination-pill therapy only | 68.83<br>(68.47 - 69.19) | 73.11<br>(72.60 - 73.63) | 61.03<br>(60.40 - 61.67) | 69.94<br>(68.26 - 71.63) |
| Difference | **3.17\*\*\***<br>**(2.79 - 3.55)** | **3.88\*\*\***<br>**(3.34 - 4.42)** | **1.76\*\*\***<br>**(1.09 - 2.44)** | **3.62\*\*\***<br>**(1.83 - 5.40)** |
| Observations | 325,600 | 174,306 | 93,122 | 13,736 |

<sup>a</sup> Logistic regression was employed to analyze medication adherence, while linear regression was used for medication possession ratios. Average marginal effects, along with 95% confidence intervals, were reported. All models were adjusted for 12 Charlson comorbidities, risk factors (alcohol use, tobacco use, obesity, and lipid disorders), gender, age groups (aged 18-34, aged 35-44, aged 45-54, and aged 55-64), race/ethnicity, and fixed effects for the year and month of the index date.
<sup>b</sup> Those who used combination-pill therapy comprise individuals who exclusively used FDC-antihypertensives during the one-year follow-up periods from the antihypertensive index date. Those who used multi-pill therapy comprise individuals who did not use FDC-antihypertensives during the one-year follow-up periods.
<sup>c</sup> The reported differences reflect the predicted outcomes, specifically the average marginal effects, for individuals receiving combination-pill therapy compared with those receiving multi-pill therapy.
\*\*\* $P < 0.001$ , \*\* $P < 0.01$ , \* $P < 0.05$

### Association between Combination-Pill Therapy and Healthcare Utilization and Costs

Compared with multi-pill therapy, those using combination-pill therapy experienced, on average, 220 fewer hypertension-related ED visits (95% CI: -235 – -204), 153 fewer hypertension-related inpatient admissions (95% CI: -160 – -146), 270 fewer CVD-related ED visits (95% CI: -287 – -254), and 181 fewer CVD-related inpatient admissions (95% CI: -189 – - 173) per 1,000 individuals one year after the index date (**Table 3**). These effects were most pronounced among NHB adults. Compared with multi-pill therapy, combination-pill therapy use among NHB adults was associated with reductions of 287 (95% CI: -318 – -256) hypertension-related ED visits, 192 (95% CI: -204 – -180) hypertension-related inpatient visits, 337 (95% CI: - 368 – -305) CVD-related ED visits, and 215 (95% CI: -228 – -202) CVD-related inpatient visits per 1,000 individuals one year after the index date. Among NHW adults, combination-pill therapy was associated with reductions of 172 (95% CI: -192 – -152) hypertension-related ED visits, 134 (95% CI: -142 – -125) hypertension-related inpatient visits, 225 (95% CI: -246 – - 204) CVD-related ED visits, and 167 (95% CI: -177 – -158) CVD-related inpatient visits per 1,000 individuals one year after the index date. Combination-pill therapy was associated with the least reduction in ED and inpatient visits among Hispanic adults.

**Table 3.**
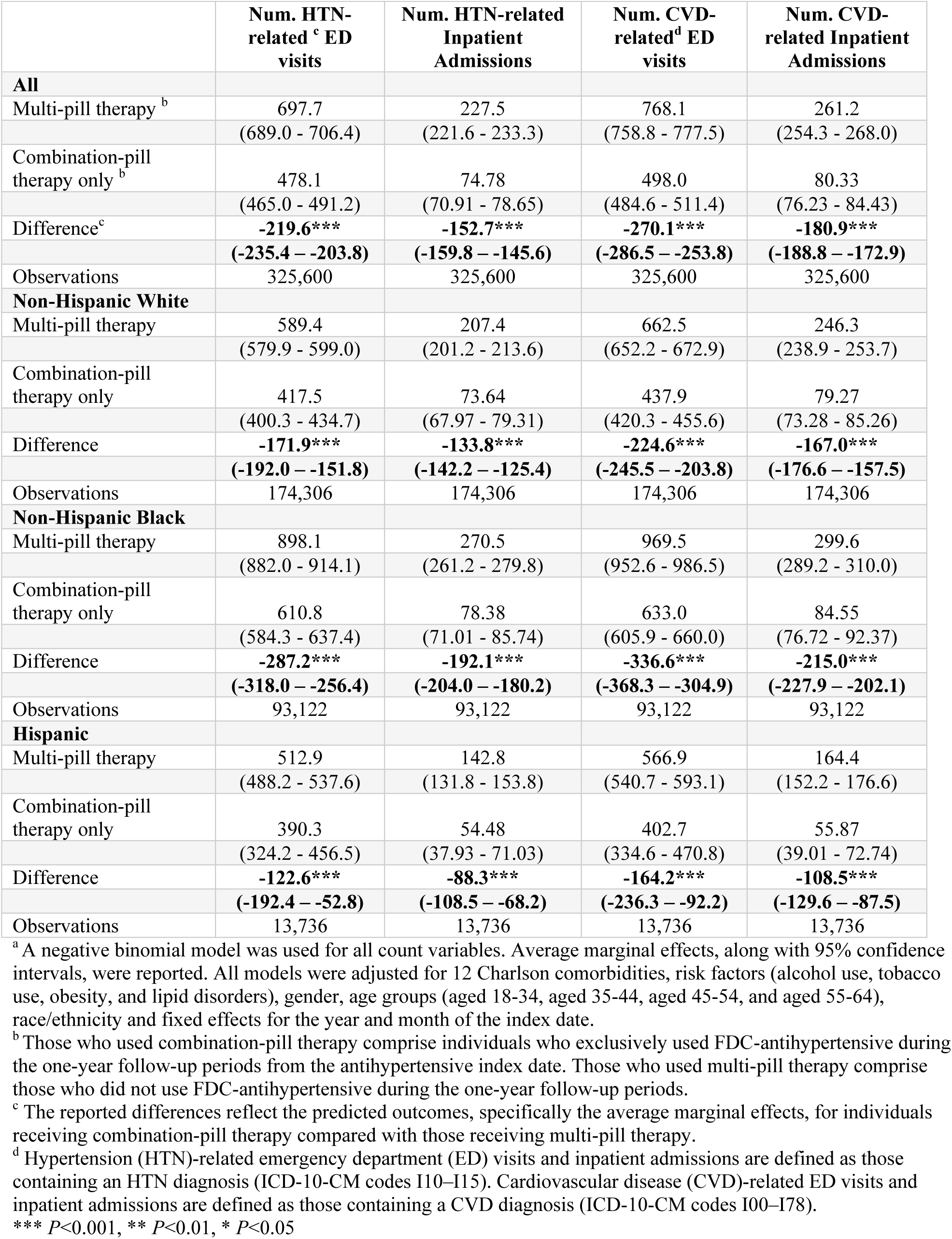
The Association of Combination-Pill Therapy Use with Hypertension- and Cardiovascular Disease-Related Health Care Utilization (per 1,000 individuals)^a^

|  | **Num. HTN-related<sup>c</sup> ED visits** | **Num. HTN-related Inpatient Admissions** | **Num. CVD-related<sup>d</sup> ED visits** | **Num. CVD-related Inpatient Admissions** |
| --- | --- | --- | --- | --- |
| **All** |  |  |  |  |
| Multi-pill therapy<sup>b</sup> | 697.7 | 227.5 | 768.1 | 261.2 |
|  | (689.0 - 706.4) | (221.6 - 233.3) | (758.8 - 777.5) | (254.3 - 268.0) |
| Combination-pill therapy only<sup>b</sup> | 478.1 | 74.78 | 498.0 | 80.33 |
|  | (465.0 - 491.2) | (70.91 - 78.65) | (484.6 - 511.4) | (76.23 - 84.43) |
| Difference<sup>c</sup> | **-219.6\*\*\*** | **-152.7\*\*\*** | **-270.1\*\*\*** | **-180.9\*\*\*** |
|  | **(-235.4 – -203.8)** | **(-159.8 – -145.6)** | **(-286.5 – -253.8)** | **(-188.8 – -172.9)** |
| Observations | 325,600 | 325,600 | 325,600 | 325,600 |
| **Non-Hispanic White** |  |  |  |  |
| Multi-pill therapy | 589.4 | 207.4 | 662.5 | 246.3 |
|  | (579.9 - 599.0) | (201.2 - 213.6) | (652.2 - 672.9) | (238.9 - 253.7) |
| Combination-pill therapy only | 417.5 | 73.64 | 437.9 | 79.27 |
|  | (400.3 - 434.7) | (67.97 - 79.31) | (420.3 - 455.6) | (73.28 - 85.26) |
| Difference | **-171.9\*\*\*** | **-133.8\*\*\*** | **-224.6\*\*\*** | **-167.0\*\*\*** |
|  | **(-192.0 – -151.8)** | **(-142.2 – -125.4)** | **(-245.5 – -203.8)** | **(-176.6 – -157.5)** |
| Observations | 174,306 | 174,306 | 174,306 | 174,306 |
| **Non-Hispanic Black** |  |  |  |  |
| Multi-pill therapy | 898.1 | 270.5 | 969.5 | 299.6 |
|  | (882.0 - 914.1) | (261.2 - 279.8) | (952.6 - 986.5) | (289.2 - 310.0) |
| Combination-pill therapy only | 610.8 | 78.38 | 633.0 | 84.55 |
|  | (584.3 - 637.4) | (71.01 - 85.74) | (605.9 - 660.0) | (76.72 - 92.37) |
| Difference | **-287.2\*\*\*** | **-192.1\*\*\*** | **-336.6\*\*\*** | **-215.0\*\*\*** |
|  | **(-318.0 – -256.4)** | **(-204.0 – -180.2)** | **(-368.3 – -304.9)** | **(-227.9 – -202.1)** |
| Observations | 93,122 | 93,122 | 93,122 | 93,122 |
| **Hispanic** |  |  |  |  |
| Multi-pill therapy | 512.9 | 142.8 | 566.9 | 164.4 |
|  | (488.2 - 537.6) | (131.8 - 153.8) | (540.7 - 593.1) | (152.2 - 176.6) |
| Combination-pill therapy only | 390.3 | 54.48 | 402.7 | 55.87 |
|  | (324.2 - 456.5) | (37.93 - 71.03) | (334.6 - 470.8) | (39.01 - 72.74) |
| Difference | **-122.6\*\*\*** | **-88.3\*\*\*** | **-164.2\*\*\*** | **-108.5\*\*\*** |
|  | **(-192.4 – -52.8)** | **(-108.5 – -68.2)** | **(-236.3 – -92.2)** | **(-129.6 – -87.5)** |
| Observations | 13,736 | 13,736 | 13,736 | 13,736 |

<sup>a</sup> A negative binomial model was used for all count variables. Average marginal effects, along with 95% confidence intervals, were reported. All models were adjusted for 12 Charlson comorbidities, risk factors (alcohol use, tobacco use, obesity, and lipid disorders), gender, age groups (aged 18-34, aged 35-44, aged 45-54, and aged 55-64), race/ethnicity and fixed effects for the year and month of the index date.
<sup>b</sup> Those who used combination-pill therapy comprise individuals who exclusively used FDC-antihypertensive during the one-year follow-up periods from the antihypertensive index date. Those who used multi-pill therapy comprise those who did not use FDC-antihypertensive during the one-year follow-up periods.
<sup>c</sup> The reported differences reflect the predicted outcomes, specifically the average marginal effects, for individuals receiving combination-pill therapy compared with those receiving multi-pill therapy.
<sup>d</sup> Hypertension (HTN)-related emergency department (ED) visits and inpatient admissions are defined as those containing an HTN diagnosis (ICD-10-CM codes I10–I15). Cardiovascular disease (CVD)-related ED visits and inpatient admissions are defined as those containing a CVD diagnosis (ICD-10-CM codes I00–I78).
\*\*\* $P < 0.001$ , \*\* $P < 0.01$ , \* $P < 0.05$

Among those with non-capitated Medicaid plans, compared with multi-pill therapy, combination-pill therapy was associated with a $2,862 per-person reduction in total hypertension-related medical costs one year after the index date (95% CI: -$3,035 – -$2,689), including $225 (95% CI: -$251 – -$199) for hypertension-related ED visits and $2,280 (95% CI: -$2,464 – -$2,096) for hypertension-related inpatient admissions (**Table 4**). Combination-pill therapy was also associated with a reduction of total CVD-related medical costs by $3,779 per person (95% CI: -$3,978 – -$3,581), with reductions of $269 (95% CI: -$295 – -$242) for CVD-related ED visits and $2,798 (95% CI: -$3,018 – -$2,578) for CVD-related inpatient visits.

**Table 4.**
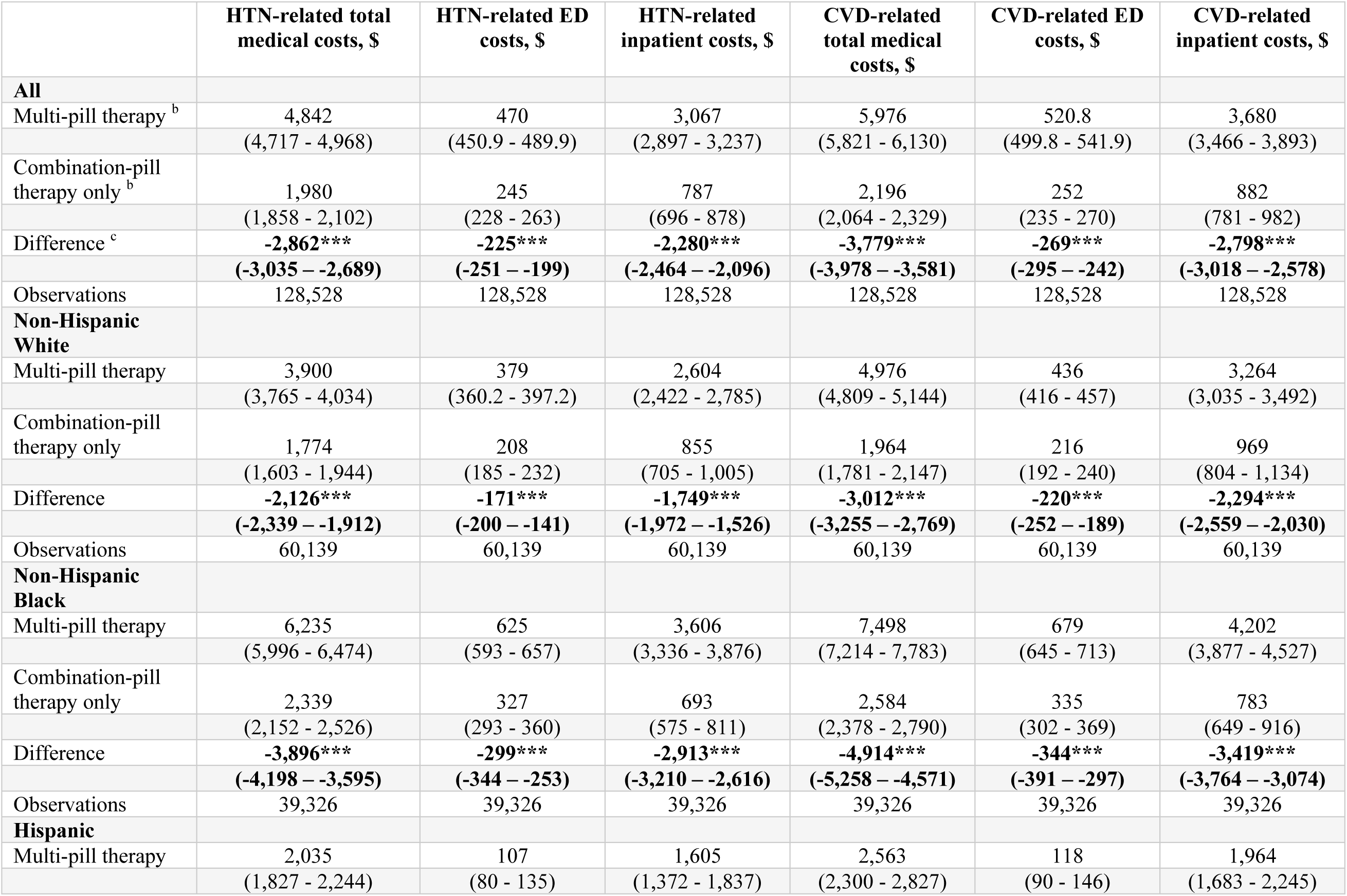
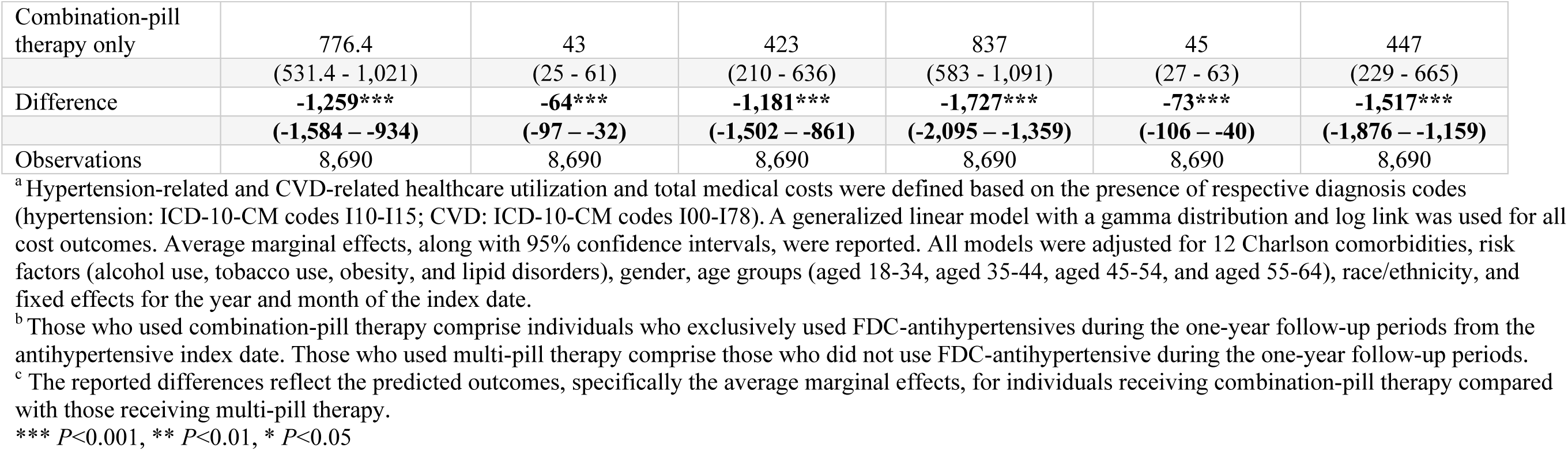
The Association of Combination-Pill Therapy Use with Medical Costs Associated with Hypertension and Cardiovascular Disease (CVD) (per individual)^a^

Among NHB adults, combination-pill therapy was associated with a reduction of hypertension-related medical costs by $3,896 per-person (95% CI: -$4,198 – -$3,595), compared with $2,126 (95% CI: -$2,339 – -$1,912) for NHW adults and $1,259 (95% CI: -$1,584 – -$934) for Hispanic adults (P<0.001). In addition, combination-pill therapy was associated with a reduction of CVD-related medical costs by $4,914 per-person (95% CI: -$5,258 – -$4,571) among NHB adults, compared with $3,012 (95% CI: -$3,255 – -$2,769) for NHW adults and $1,727 (95% CI: - $2,095 – -$1,359) for Hispanic adults (p<0.001).

### Sensitivity Analyses

In sensitivity analyses, compared with multi-pill therapy, combination-pill therapy was associated with a significant reduction in all-cause healthcare utilization and medical costs one year after the index date, with a stronger association observed among NHB adults (p<0.001, **Appendix Table 3 and 4**). Further, 82,126 (22%) used both combination- and multi-pill therapy during the same year, (**Appendix Table 5**). Including individuals who used both therapies did not alter the findings or conclusions (**Appendix Figure 3**, **Appendix Table 6-9**).

### Comparison of Annual and Out-Of-Pocket Costs between Combination-and Multi-Pill Therapy groups

The annual per-person costs for combination-pill therapy ($111 95% CI: $107 – $115) were significantly lower than those for multi-pill therapy ($273, 95% CI: $270 – $275). The patient OOP costs for combination-pill therapy ($8) were also lower than those for multi-pill therapy ($13) (P<0.001) (**Figure 1**). However, among those who used combination-pill therapy, NHB beneficiaries had similar annual medication costs ($110) to NHW ($107) but significantly higher than Hispanic adults ($61).

**Figure 1.**
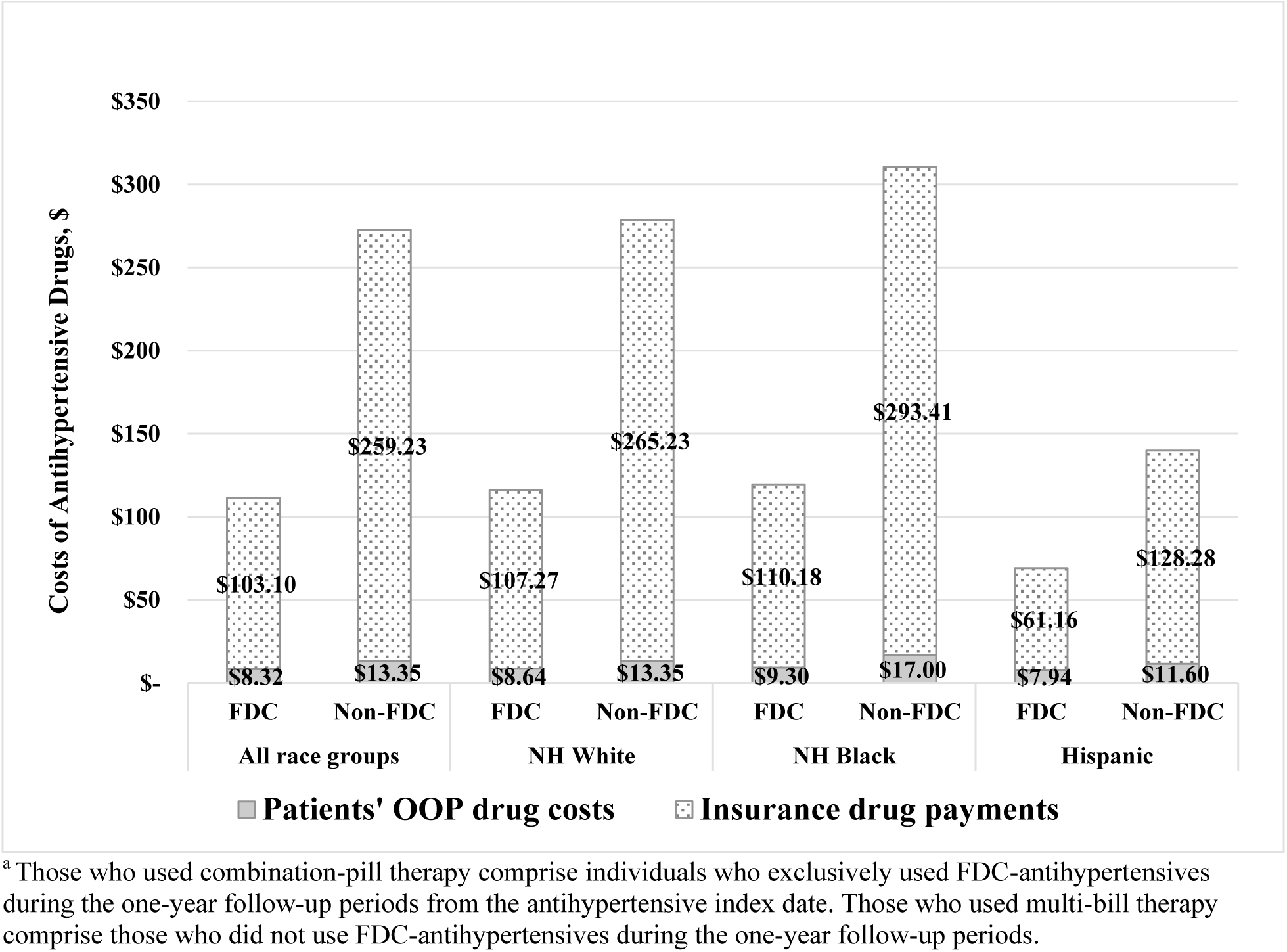
Insurance, and Patients’ Out-of-Pocket Costs for Combination-Pill Therapy and Multi-Pill Therapy by Racial and Ethnic Groups, MarketScan Medicaid, 2016-2022^a^. ^a^ Those who used combination-pill therapy comprise individuals who exclusively used FDC-antihypertensives during the one-year follow-up periods from the antihypertensive index date. Those who used multi-bill therapy comprise those who did not use FDC-antihypertensives during the one-year follow-up periods.

## Discussion

In this cohort study of Medicaid beneficiaries with hypertension, we found that 9% used combination-pill therapy to manage their blood pressure. Combination-pill therapy was significantly associated with higher medication adherence, fewer ED visits and hospitalizations, and lower medical costs compared with multi-pill therapy. Combination-pill therapy was also associated with lower Medicaid payments and patient OOP costs than multi-pill therapy.

However, improvement in medication adherence was smaller among NHB adults than NHW adults for combination-pill therapy versus multi-pill therapy, but NHB adults showed a more pronounced association with reduced healthcare utilization and costs.

Compared with the reported prevalence of FDC-antihypertensive/combination-pill therapy among all U.S. adults with hypertension based on National Health and Nutrition Examination Survey (NHANES) 2017-March 2020 (27.8%),^9^ we found a lower prevalence of Medicaid beneficiaries using FDC-antihypertensives exclusively (9%, our main analysis definition of combination-pill therapy) or using a mix of FDC-antihypertensives and other antihypertensives during the same year (22%; our more inclusive sensitivity analysis definition of combination-pill therapy) between 2017-2021. Previous reports indicated a decrease in FDC-antihypertensive use from 2012 to 2020,^9^ with Medicare and Medicaid FDC claims decreasing from 2016 to 2020.^29^ The majority of FDC-antihypertensives are available in generic form,^29^ and our analysis showed that Medicaid payments and patient OOP costs were lower for combination-pill therapy. Given the small proportion of Medicaid beneficiaries with hypertension using combination-pill therapy, barriers to use of combination-pill therapy may not be attributed to costs. Prior work has documented barriers including lack of sufficiently diverse FDC-antihypertensive therapeutic options with respect to medication classes and dosage; one study estimated that nearly two-thirds of US adults with hypertension were using a regimen not available as an FDC-antihypertensive product. ^11^

Consistent with the literature,^30,31^ our analysis demonstrated that combination-pill therapy was associated with significantly higher medication adherence. However, combination-pill therapy was less associated with medication adherence in NHB adults (1.76, 95% CI: 1.09 - 2.44) compared with NHW adults (3.88, 95% CI: 3.34 - 4.42). This finding may be attributable to several reasons. First, the majority of FDC-antihypertensives were combinations of ACE inhibitors or ARBs plus thiazide diuretics,^29^ and ACE inhibitors and ARBs may be insufficient for improving hypertension control for NHB adults, possibly due to poorer medication adherence and/or differences in physiological pathways.^32–34^ The AHA/ACC guideline recommends initial antihypertensive treatment with a thiazide diuretic or CCBs for NHB adults. However, there are no CCB/thiazide diuretic FDC-antihypertensives, which represents a gap for guideline-recommended combination-pill therapy for NHB adults, who may have a less robust response to ACEIs, ARBs, and beta-blockers.^35^ Second, there may be differences in plan enrollment and coverage formularies at the state or plan level, particularly in states with managed care organizations. As shown in our analysis, there were some slight variations in costs for NHB beneficiaries compared with NHW and Hispanic beneficiaries. NHB beneficiaries may be more likely to be enrolled in plans with higher cost-sharing responsibilities compared with NHW and Hispanic Medicaid beneficiaries.^36^ However, our data did not include data on insurance benefit designs, so this is beyond the scope of the present analysis and may be investigated in national Medicaid claims databases in the future.

We observed that combination-pill therapy was associated with greater reductions in ED and inpatient visits among NHB adults compared with NHW adults, despite a smaller association with medication adherence. This could be because NHB adults initially had much lower medication adherence and higher healthcare utilization and costs. Thus, they might benefit more from using combination-pill therapy. If NHB adults had medication adherence comparable to NHW adults, the reduction in healthcare utilization and costs might have been even larger. In contrast, Hispanic adults had higher medication adherence than NHB adults initially, but the lowest healthcare utilization and costs. Previous studies have reported that Hispanic adults are less likely to seek health care due to language, cultural barriers, and immigration status.^37^ This could partially explain why the associations between combination-pill therapy and lower healthcare utilization and costs were smaller among Hispanic adults compared with NHW adults.

Our study has important implications. While combination-pill therapy has the potential to improve medication adherence, the available options in the US may not offer the variety of class and dose combinations preferred by clinicians. An analysis of 2015-2020 NHANES data found 189 unique therapeutic class combination regimens among US adults using antihypertensives, only 7 of which (3.7%) were available as FDC-antihypertensives.^11^ Similar findings are seen among combination regimens in the Systolic Blood Pressure Intervention Trial (SPRINT).^11,38^ There are no available combinations containing four antihypertensives, which were used by over 25% of SPRINT participants. The availability of only two- or three-drug FDC-antihypertensives, and no four- or five-drug FDC-antihypertensives, limits the treatment options for patients in the U.S. who require multiple antihypertensives to achieve hypertension control.^39^ Furthermore, the need for triple and quadruple drug combinations may increase because ACC/AHA hypertension treatment guidelines have recommended more intensive treatment goals (lower target blood pressures).^40^ Factors such as drug shortages, changes in insurance coverage or cost-sharing tiers, or concerns about side effects may deter initiation or prompt switching from FDC-antihypertensives to other antihypertensives. Moreover, while 90% of Medicaid plans cover at least four FDC-antihypertensives, and 100% cover at least one FDC-antihypertensive, individual drug coverage and costs vary by state Medicaid programs.^39^ Some drugs are on preferred (Tier 1) or non-preferred (Tier 2) tiers, which may influence clinicians’ prescribing behavior. To address these gaps, policymakers and pharmaceutical companies could consider exploring and broadening FDC-antihypertensive options with a wider range of drug class combinations to suit the variety of treatment needs faced by prescribing clinicians.^41^

### Limitations

We acknowledge several limitations. First, we only tracked short-term outcomes over a single year of combination-pill therapy use, leaving the long-term impacts of combination-pill therapy, such as mortality and productivity, unknown. Second, as we used a multistate Medicaid claims database, our findings may not be generalizable to individuals covered by other types of insurance, particularly because the price of FDC-antihypertensives covered by Medicaid tends to be lower than that of equivalent drugs covered by Medicare and private insurance.^29^ Additionally, individual drug coverage and costs vary by state Medicaid programs, which may lead to disparities in access and treatment patterns across different states. However, our data do not include state identifiers, preventing us from differentiating Medicaid medication coverage policies or decisions at the state level. Third, our claims database only shows whether prescriptions were filled and picked up, not if patients actually took the medications. In addition, we could not analyze data at the prescribing clinician level, accounting for variance due to clinician behavior. These issues may lead to potentially overestimation of medication adherence. Fourth, our main analysis focused on individuals exclusively using combination-pill therapy (excluding those who used both combination-pill therapy and other antihypertensives in a single year), which may limit generalizability. However, our sensitivity analyses included individuals who used both combination- and multi-pill therapy in the same year, and results were consistent with the main findings. Finally, we could not ascertain the doses of FDC-antihypertensives, which may affect the healthcare utilization outcomes we assessed.

## Conclusions

This large-scale study among Medicaid beneficiaries in multiple states (5 to 8 states from year to year) across the US found that combination-pill therapy was associated with higher medication adherence, lower healthcare utilization, and lower healthcare costs for the Medicaid program and beneficiaries. However, the magnitude of these associations differs by racial and ethnic groups. These research findings may help inform policies related to FDC-antihypertensive medication coverage or formulary decisions, as well as programs aimed at improving medication adherence and hypertension control. In addition, our findings can provide important evidence for clinicians when selecting antihypertensive medications for patients.

## Data Availability

Data were obtained from the MerativeTM MarketScan® multistate Medicaid claims database from 2016 to 2022.

## Acknowledgments

The authors are genuinely grateful to Michael Schooley (Centers for Disease Control and Prevention) for their guidance, suggestions, and article review.

